# Genomic characterization of upper urinary tract urothelial carcinoma and clonal evolution of intravesical recurrences

**DOI:** 10.64898/2026.02.18.26346396

**Authors:** J. Alberto Nakauma-Gonzalez, Veronika Bahlinger, Thomas van Doeveren, Harmen J.G. van de Werken, Jozien Helleman, Justine Pasanisi, Julien Masliah-Planchon, Ivan Bieche, Therese Wilhelm, Geert J.L.H. van Leenders, Maria Fernanda Lara, Francisco Porcel-Pastrana, Enrique Gómez-Gómez, Raúl M. Luque, Lucia García-Morales, Markus Eckstein, Robert Stöhr, Danijel Sikic, Isabel García Muñoz, Juan Daniel Prieto Cuadra, Maria José Lozano, Martina Álvarez, Elisa Matas-Rico, Arndt Hartmann, Bernardo Herrera-Imbroda, Yves Allory, Joost L. Boormans

**Affiliations:** Department of Urology, Erasmus MC Cancer Institute, University Medical Center Rotterdam, Rotterdam 3015 GD, the Netherlands; Institute of Pathology, University Hospital Erlangen-Nürnberg, Friedrich-Alexander-Universität Erlangen-Nürnberg; Department of Pathology and Neuropathology, University Hospital and Comprehensive Cancer Center Tuebingen, Tuebingen, Germany; Comprehensive Cancer Center Erlangen-EMN (CCC ER-EMN), Erlangen, Germany; Department of Immunology, Erasmus MC Cancer Institute, University Medical Center Rotterdam, Rotterdam 3015 GD, the Netherlands; Department of Genetics, Institut Curie, PSL University, 26 rue d’Ulm, 75005, Paris, France; Department of Pathology, Erasmus MC Cancer Institute, University Medical Center Rotterdam, Rotterdam 3015 GD, the Netherlands; Plataforma en Nanomedicina-IBIMA-Bionand; Reseach Group in Maternal-Fetal Medicine, Epigenetics, Womeńs Diseases and Reproducitve Healht. Dpt. of Surgical Specialties, Biochemistry and Immunology; OBGYN Area; Málaga University. 2907, Málaga, Spain; Maimonides Institute for Biomedical Research of Córdoba (IMIBIC), 14004 Cordoba, Spain; Urology department, Reina Sofía University Hospital, University of Córdoba, Córdoba, Spain; Department of Cell Biology, Physiology, and Immunology, University of Cordoba; CIBER Fisiopatologia de la Obesidad y Nutricion (CIBEROBN); 14014, Córdoba, Spain; Department of Urology and Pediatric Urology, Friedrich-Alexander-University Erlangen-Nuremberg, Erlangen, Germany; Servicio de Anatomía Patológica. Hospital Universitario Virgen de la Victoria, Málaga, Spain; IBIMA Plataforma Bionand, Instituto de Investigación Biomédica de Málaga. Grupo de Anatomía Patológica, Málaga, Spain; Departamento de Fisiología Humana, Histología Humana, Anatomía Patología y Educación Física y del Deporte. Facultad de Medicina, Universidad de Málaga, Málaga, Spain; IBIMA Plataforma Bionand, Instituto de Investigación Biomédica de Málaga. Grupo de Investigación Clínica y Traslacional en Cáncer, Málaga, Spain; Laboratorio de Biología Molecular del Cáncer. Centro de Investigaciones Médico-Sanitarias. Universidad de Málaga, Málaga, Spain; IBIMA Plataforma Bionand, Instituto de Investigación Biomédica de Málaga. Grupo de investigación traslacional en Oncología genitourinaria, Malaga, Spain; Departamento de Biología Celular, Genética y Fisiología, Universidad de Málaga, Spain; Servicio de Urología. Hospital Universitario Virgen de la Victoria, Málaga, Spain; Université Paris-Saclay, UVSQ, Institut Curie, Department of Pathology, Saint-Cloud, France

**Keywords:** Clonal evolution, Diagnostic markers, Genomic subtypes, Intravesical recurrence, Targeted DNA sequencing, Upper tract urothelial cancer

## Abstract

**Background and Objective:** Patients with upper urinary tract urothelial carcinoma (UTUC) undergoing radical surgery are at high risk of developing intravesical recurrences (IVR). Molecular correlates of IVR after UTUC surgery are poorly understood, while therapeutic targets for UTUC and diagnostic markers for IVR are lacking. To address these needs, we investigated the genomic landscape of UTUC and paired IVR samples.

**Methods:** We performed targeted DNA-sequencing of 571 genes in a multi-institutional international cohort of 414 UTUC patients who received radical surgery. Clonality and evolution were assessed in 79 UTUC paired with 104 IVR.

**Key Findings and Limitations:** The mutually exclusive mutational profile of 414 UTUC samples revealed five genomic subtypes with distinct clinicopathological and molecular characteristics, and potential vulnerabilities to targeted therapy. Surgical intervention, specifically diagnostic ureteroscopy, showed a stronger association with IVR than genomic subtypes or individual genes. Clonal evolution occurred in 92% of UTUC-IVR cases via four evolutionary paths with mutations in *FGFR3*, *HRAS* and the *TERT* promoter as potential markers for surveillance after surgery. Limitations include cohort heterogeneity and the selected gene-targeted sequencing approach.

**Conclusions and Clinical Implications:** Our findings resolve the genomic heterogeneity of UTUC and its clonal relationship with IVR, providing a framework for molecularly guided surveillance and targeted therapeutic strategies for UTUC.

## Introduction

Urothelial carcinoma (UC), with over 600,000 new diagnoses every year, is among the most common cancers worldwide [1,2]. Upper tract UC (UTUC) is less common than UC of the bladder (UCB), accounting for only 5-10% of all diagnosed UCs [3]. Due to its relatively low incidence, UTUC is understudied, resulting in a poorly understood biology and suboptimal treatment strategies [4–6]. Furthermore, up to 60% of UTUC patients have invasive disease at diagnosis *versus* only 25% in UCB, impacting survival probabilities [7]. At the genomic level, UTUC is associated with Lynch syndrome, and *FGFR3* and *HRAS* mutations are more frequent than in UCB [8–10].

Guideline-recommended treatment for nonmetastatic high-risk UTUC patients is radical nephroureterectomy (RNU) [11]. Between 22-47% of patients undergoing RNU develop intravesical recurrence (IVR) within two years after surgery [12]. Known risk factors for IVR in UTUC patients treated with RNU are: a history of UCB [13,14], tumor location in the distal ureter, multifocality of the primary UTUC, and high-grade disease [12,15]. Based on clinical observations, the seeding hypothesis has been proposed as a mechanism of IVR after RNU [16]. Cancer cells from the UTUC detach during surgery, spread via the ureter to the bladder, where they give rise to IVRs [17]. Genomic studies have reported that most UTUC and IVR share a common ancestor, supporting the seeding hypothesis [18,19]. However, these studies relied on small cohorts of ≤16 patients, stressing the need for validation in larger cohorts.

There is a clear knowledge gap on the molecular biology of UTUC and the high risk of IVR after RNU. To monitor possible IVR in UTUC, patients undergo close surveillance of the bladder by invasive cystoscopy, which is uncomfortable and a psychological burden to patients. Therefore, it is crucial to clarify the clonal seeding hypothesis as the main mechanism of IVR following UTUC to identify molecular markers for surveillance. To address these needs and the lack of therapeutic molecular targets for UTUC, we performed a genomic analysis in 518 tumor samples (414 UTUC + 108 IVR) derived from 414 UTUC patients and sought to clarify the clonal relationship between UTUC and IVR, as well as molecular correlates and evolution of IVR.

## Patients and methods

The Supplementary material provides full details of the methods.

### Samples and patient cohort characteristics

A multi-institutional international cohort of 414 UTUC patients (30.2% females) who underwent radical surgery between 2002 and 2020 was investigated. UTUC samples plus 104 IVR samples from 79 patients (16 had multiple IVRs) were retrospectively collected (276 patients; central pathologically reviewed) or in the context of a prospective clinical trial (138 patients; REBACARE trial, METC 2017-227, NL60919.078.17) [20]. The median age and follow-up were 72 years (quartile 1-3 (Q1-Q3): 65-77) and 24.8 months (Q1-Q3: 16.0-65.1), respectively. Among other differences (**Supplementary Table 1**), the median follow-up was shorter in the prospective (23.6 months, Q1-Q3: 21.2-25.4) than in the retrospective cohorts (41.2 months, Q1-Q3: 11.9-83.5; p=0.0001, two-sided Wilcoxon rank-sum test), but IVR-free survival was comparable between the two cohort groups (**Supplementary Fig. 1**). Of 414 UTUC patients, 47 (11.4%) had a history of UCB (UCB-UTUC) and 299 (72.2%) had primary UTUC, of whom 87 (21.0%) developed an IVR (UTUC-IVR) after UTUC surgery. In 68 cases (16.4%), IVR or history of UCB was unknown. The follow-up time in the UTUC-IVR subgroup was 24.6 months (Q1-Q3: 21.6-51.1) and comparable to the rest of the cohort with 25.2 months (Q1-Q2: 16.0-66.3). The median time to IVR was 12.2 months (Q1-Q3: 5.7-19.1), and 78.6% occurred within two years after surgery. UTUC in the renal pelvis was more frequent (n=180; 43.5%) than in the ureter (n=118; 28.5%). In 15 cases (3.6%), tumors in the renal pelvis and ureter (multilocal) were detected simultaneously at diagnosis, while tumor location was unknown for 101 cases (24.4%). In 69.6% of cases, UTUC presented high-grade disease, and concomitant carcinoma *in situ* (CIS) was identified in 29 (7.0%) patients. A summary of the cohort is presented in **Table 1**.

**Table 1.** Clinicopathological characteristics of 414 patients with upper tract urothelial carcinoma who received radical surgery stratified by a history of urothelial carcinoma of the bladder or subsequent intravesical recurrence.

| Cohort characteristics | Total cohort<br>(N=414) | UTUC subgroups (N=346) |  |  | P |
| --- | --- | --- | --- | --- | --- |
|  |  | UCB-UTUC<br>(N=47) | UTUC-IVR<br>(N=87) | UTUC-only<br>(N=212) |  |
| Patient Institute, n (%) |  |  |  |  |  |
| ERL | 161 (38.9) | 0 (0) | 21 (24.1) | 74 (34.9) | <0.0001 |
| ERA* | 138 (33.3) | 0 (0) | 42 (48.3) | 96 (45.3) |  |
| MAL | 96 (23.2) | 28 (59.6) | 24 (27.6) | 42 (19.9) |  |
| EME | 19 (4.6) | 19 (40.4) | 0 (0) | 0 (0) |  |
| Age |  |  |  |  |  |
| Median, years (Q1-Q3) | 72 (65-77) | 73 (61-78) | 71 (66-75) | 72 (65-77) | 0.90** |
| Unknown, n (%) | 4 (1.0) | 0 (0) | 0 (0) | 0 (0) |  |
| Sex, n (%) |  |  |  |  |  |
| Female | 125 (30.2) | 9 (19.1) | 29 (33.3) | 65 (30.7) | 0.18 |
| Male | 287 (69.3) | 38 (80.9) | 58 (66.7) | 147 (69.3) |  |
| Unknown | 2 (0.5) | 0 (0) | 0 (0) | 0 (0) |  |
| Follow up |  |  |  |  |  |
| Median, months (Q1-Q3) | 25 (16-65) | 64 (43-90) | 25 (22-51) | 24 (14-45) | <0.0001 |
| Unknown, n (%) | 21 (5.1) | 0 (0) | 2 (2.5) | 5 (2.5) |  |
| Location, n (%) |  |  |  |  |  |
| Renal pelvis | 180 (43.5) | 19 (40.4) | 33 (37.9) | 92 (43.4) | <0.0001<br>(0.03***) |
| Ureter | 118 (28.5) | 20 (42.6) | 32 (36.8) | 46 (21.7) |  |
| Multilocal | 15 (3.6) | 8 (17.0) | 1 (1.1) | 0 (0) |  |
| Unknown | 101 (24.4) | 0 (0) | 21 (24.1) | 74 (34.9) |  |
| N stage, n (%) |  |  |  |  |  |
| N0 | 239 (57.7) | 14 (29.8) | 61 (70.1) | 144 (67.9) | 0.27 |
| N1 | 26 (6.3) | 1 (2.1) | 4 (4.6) | 12 (5.7) |  |
| N2 | 11 (2.7) | 0 (0) | 0 (0) | 10 (4.7) |  |
| NX or unknown | 138 (33.3) | 32 (68.1) | 22 (25.3) | 46 (21.7) |  |
| Surgery, n (%) |  |  |  |  |  |
| RNU | 380 (92.1) | 39 (83.0) | 76 (87.7) | 199 (93.9) | 0.030 |
| Partial ureterectomy | 32 (7.4) | 8 (17.0) | 11 (12.3) | 13 (6.1) |  |
| Not specified | 2 (0.5) | 0 (0) | 0 (0) | 0 (0) |  |
| Preoperative URS, n (%) |  |  |  |  |  |
| Yes | 94 (22.7) | 11 (23.4) | 37 (42.5) | 46 (21.7) | 0.0002 |
| No | 85 (20.5) | 20 (42.6) | 11 (12.6) | 54 (25.5) |  |
| Unknown | 235 (56.8) | 16 (34.0) | 39 (44.8) | 112 (52.8) |  |
| Ureteral clipping, n (%) |  |  |  |  |  |
| Yes | 157 (37.9) | 22 (46.8) | 42 (48.3) | 93 (43.9) | 0.059 |
| No | 36 (8.7) | 11 (23.4) | 8 (9.2) | 17 (8.0) |  |
| Unknown | 221 (53.4) | 14 (29.8) | 37 (42.5) | 102 (48.1) |  |
| Bladder cuff excision, n (%) |  |  |  |  |  |
| No | 43 (10.4) | 12 (25.5) | 14 (16.1) | 17 (8.0) | <0.0001 |
| Yes, endoscopic | 54 (13.0) | 18 (38.3) | 9 (10.3) | 27 (12.7) | (0.046****) |
| Yes, extravesical | 76 (18.4) | 10 (21.3) | 24 (27.6) | 42 (19.8) |  |
| Yes, transvesical | 58 (14.0) | 0 (0) | 17 (19.5) | 41 (19.3) |  |
| Unknown | 183 (44.2) | 7 (14.9) | 23 (26.4) | 85 (40.1) |  |
| T stage, n (%) |  |  |  |  |  |
| Tis | 6 (1.4) | 2 (4.3) | 2 (2.2) | 2 (0.9) |  |
| Ta | 93 (22.5) | 3 (6.4) | 26 (29.9) | 52 (24.5) | <0.0001 |
| T1 | 88 (21.3) | 26 (55.3) | 19 (21.8) | 32 (15.1) |  |
| T2 | 66 (15.9) | 9 (19.1) | 15 (17.2) | 32 (15.1) |  |
| T3 | 132 (31.9) | 6 (12.8) | 22 (25.3) | 77 (36.3) |  |
| T4 | 25 (6.0) | 1 (2.1) | 3 (3.4) | 17 (8.0) |  |
| Unknown | 4 (1.0) | 0 (0) | 0 (0.0) | 0 (0) |  |
| Concomitant CIS, n (%) |  |  |  |  |  |
| Yes | 29 (7.0) | 4 (8.5) | 9 (10.3) | 16 (7.5) | 0.69 |
| No | 220 (53.1) | 43 (91.5) | 56 (64.4) | 121 (57.1) |  |
| Unknown | 165 (39.9) | 0 (0) | 22 (25.3) | 75 (35.4) |  |
| Lymphovascular invasion, n (%) |  |  |  |  |  |
| Yes | 55 (13.3) | 6 (12.8) | 9 (10.3) | 40 (18.9) | 0.008 |
| No | 184 (44.4) | 41 (87.2) | 53 (60.9) | 90 (42.4) |  |
| Unknown | 175 (42.3) | 0 (0) | 25 (28.7) | 82 (38.7) |  |
| Necrosis, n (%) |  |  |  |  |  |
| Yes | 15 (3.6) | 2 (4.3) | 3 (3.4) | 10 (4.7) | 0.37 |
| No | 172 (41.5) | 45 (95.7) | 44 (50.6) | 83 (39.2) |  |
| Unknown | 227 (54.8) | 0 (0) | 40 (46.0) | 119 (56.1) |  |
| Surgical margins, n (%) |  |  |  |  |  |
| Positive | 27 (6.5) | 7 (14.9) | 9 (10.3) | 11 (5.2) | 0.051 |
| Negative | 383 (92.5) | 40 (85.1) | 78 (89.7) | 199 (93.9) |  |
| Unknown | 4 (1.0) | 0 (0) | 0 (0) | 2 (0.9) |  |
| IVR, n (%) |  |  |  |  |  |
| Yes | 130 (31.4) | 43 (91.5) | 87 (100) | 0 (0) |  |
| No | 216 (52.2) | 4 (8.5) | 0 (0) | 212 (100) |  |
| Unknown | 68 (16.4) | 0 (0) | 0 (0) | 0 (0) |  |
| Survival status, n (%) |  |  |  |  |  |
| Alive | 248 (59.9) | 21 (44.7) | 57 (65.5) | 121 (57.1) | 0.053 |
| Death | 158 (38.2) | 26 (55.3) | 29 (33.3) | 88 (41.5) |  |
| Unknown | 8 (1.9) | 0 (0) | 1 (1.1) | 3 (1.4) |  |

UTUC: Upper tract urothelial carcinoma.
UCB-UTUC: UTUC with a previous history of UCB.
UTUC-IVR: UTUC with subsequent IVR without previous UCB.
UTUC-only: UTUC had neither previous UCB nor IVR.
ERL: patients from the University Hospital Erlangen-Nürnberg, Germany
ERA: patients from the Erasmus University Medical Center Rotterdam, the Netherlands.
MAL: patients from the Virgen de la Victoria University Hospital Malaga, Spain
EME: patients from the University of Malaga and Córdoba Biobank (Reina Sofia University Hospital), funded by the EMERGIA program, Spain.
CIS: carcinoma *in situ*.
URS: ureteroscopy
Estimates were given as median (quartile 1 (Q1) - quartile 3(Q3)) or frequency (percentage).
\* Prospective cohort.

### DNA-sequencing and clonal evolution

DNA from the tumor and matched normal were sequenced using a panel of 571 genes [21]. Somatic mutations and copy number alterations were assessed as previously described [21,22].

Clonality assessment was performed on UTUC and matched IVR using the clonal relatedness test [23,24]. For clonally related samples, changes in mutation allele frequency were used as *a proxy* for selection (selection score) to define evolutionary paths by hierarchical clustering [25].

### Statistical analysis

Analyses were performed using R v4.4.1 [26]. The clonality test was applied with Clonality v1.47.0, which is based on the Neyman-Pearson Lemma test and the conditional maximum likelihood estimate. The χ^2^ and Fisher’s exact tests were used for comparison of categorical values between groups. The Wilcoxon rank-sum test, signed-rank test, and the Kruskal-Wallis test by ranks were used to compare groups with continuous variables. Time from surgery to IVR and death was compared between groups by Kaplan-Meier estimates with the log-rank test. Patients who were lost to follow-up were censored at the date of the last cystoscopy. The Cox proportional hazards regression was applied for univariate and multivariate analysis. *P* values were adjusted for multiple testing using the Benjamini–Hochberg method.

## Results

### Clinicopathological correlates with intravesical recurrences

Time-to-event univariate Cox regression analysis (**Supplementary Fig. 2**) identified preoperative ureteroscopy (hazard ratio (HR) = 3.1, 95% confidence interval (CI): 1.7-5.8, p=0.0004), a history of UCB (HR=2.6, CI: 1.5-4.5), p=0.0007), multilocal UTUC (HR=5.1, CI: 1.6-16.3, p=0.0065) and tumor location in the ureter (HR=1.9, CI: 1.2-3.1, p=0.008) to be associated with IVR. Only bladder cuff excision was associated with reduced risk of IVR (HR=0.48, CI: 0.27-0.84, p=0.01). After applying multivariate Cox regression analysis to these variables, only preoperative ureteroscopy (HR=2.95, CI: 1.5−5.8, p=0.0017) and a history of UCB (HR=2.4, CI: 1.06-5.6, p=0.036) remained significant. Other clinicopathological variables, including positive surgical margin, CIS, high tumor grade and stage, age, sex, necrosis, lymphovascular invasion and ureteral clipping during RNU, showed no significant associations with risk of IVR.

Excluding unknowns, the UTUC-only subgroup (no previous UCB and no IVR at follow-up; n=212) represented 61.3% of patients (**Fig. 1A)** *versus* 13.6% UCB-UTUC and 25.1% UTUC-IVR. Given the association of a history of UCB with IVR, in which 91.5% of UCB-UTUC developed an IVR, both UCB-UTUC and UTUC-IVR subgroups represent patients with a high risk of IVR (**Supplementary Fig. 3**) with potentially distinct biology and characteristics leading to IVR (**Table 1**). Multilocal tumors (p<0.0001, χ^2^ test) were overrepresented in the UCB-UTUC subgroup, while age, sex, N stage, CIS, positive surgical margins, ureteral clipping during RNU, tumor necrosis and survival status were similar across UTUC subgroups. Preoperative ureteroscopy was more common in the UTUC-IVR subgroup (p=0.0002, χ^2^ test), while UTUC-only had a higher proportion of lymphovascular invasion (p=0.008, χ^2^ test) and tumors from the renal pelvis (p=0.016, χ^2^ test). Ta/Tis tumors were underrepresented in the UCB-UTUC subgroup (10.7%), compared to UTUC-IVR (32.1%) and UTUC-only (24.4%; p<0.0001, χ^2^ test). Follow-up time was comparable between UTUC-IVR (25 months; Q1-Q3: 22-51) and UTUC-only (24 months; Q1-Q3: 14-45), but it was longer in the UCB-UTUC subgroup (64 months; Q1-Q3: 43-90). Despite these differences, overall survival was comparable among the three subgroups (**Fig. 1B**).

**Figure 1.**
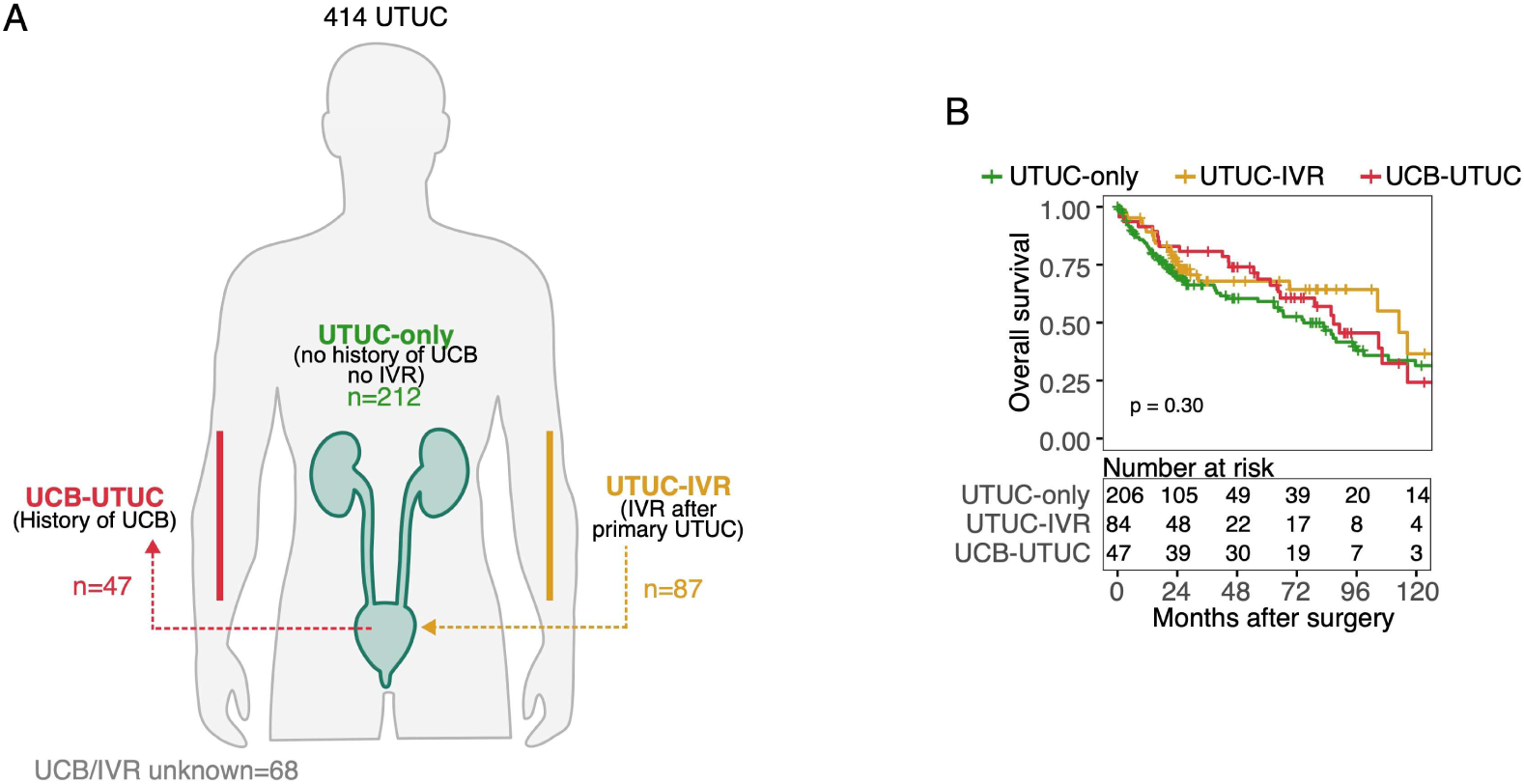
Cohort overview of 414 patients with upper urinary tract urothelial carcinoma stratified according to history of urothelial carcinoma of the bladder or intravesical recurrence. **A)** Schematic representation of all upper urinary tract urothelial carcinoma (UTUC) samples grouped according to a history of urothelial carcinoma of the bladder (UCB; UCB-UTUC), intravesical recurrence (IVR; UTUC-IVR) or a history without UCB nor IVR (UTUC-only). **B)** Kaplan–Meier curves for estimates of overall survival among UTUC subgroups. The log-rank test was used to compare the Kaplan–Meier survival curves.

### The mutational landscape of UTUC and genomic correlates with intravesical recurrences

The median tumor mutational burden (TMB) of UTUC samples was 4.18 (Q1-Q3: 2.79-6.62) mutations per megabase-pair (**Fig. 2A**), with the lowest TMB in the UTUC-only subgroup (p=0.032, two-sided Wilcoxon rank-sum test). Although UTUC location might be associated with IVR risk (**Supplementary Fig. 2; Supplementary Table 2**), no difference in TMB was observed between the renal pelvis and ureter (**Fig. 2A**).

**Figure 2.**
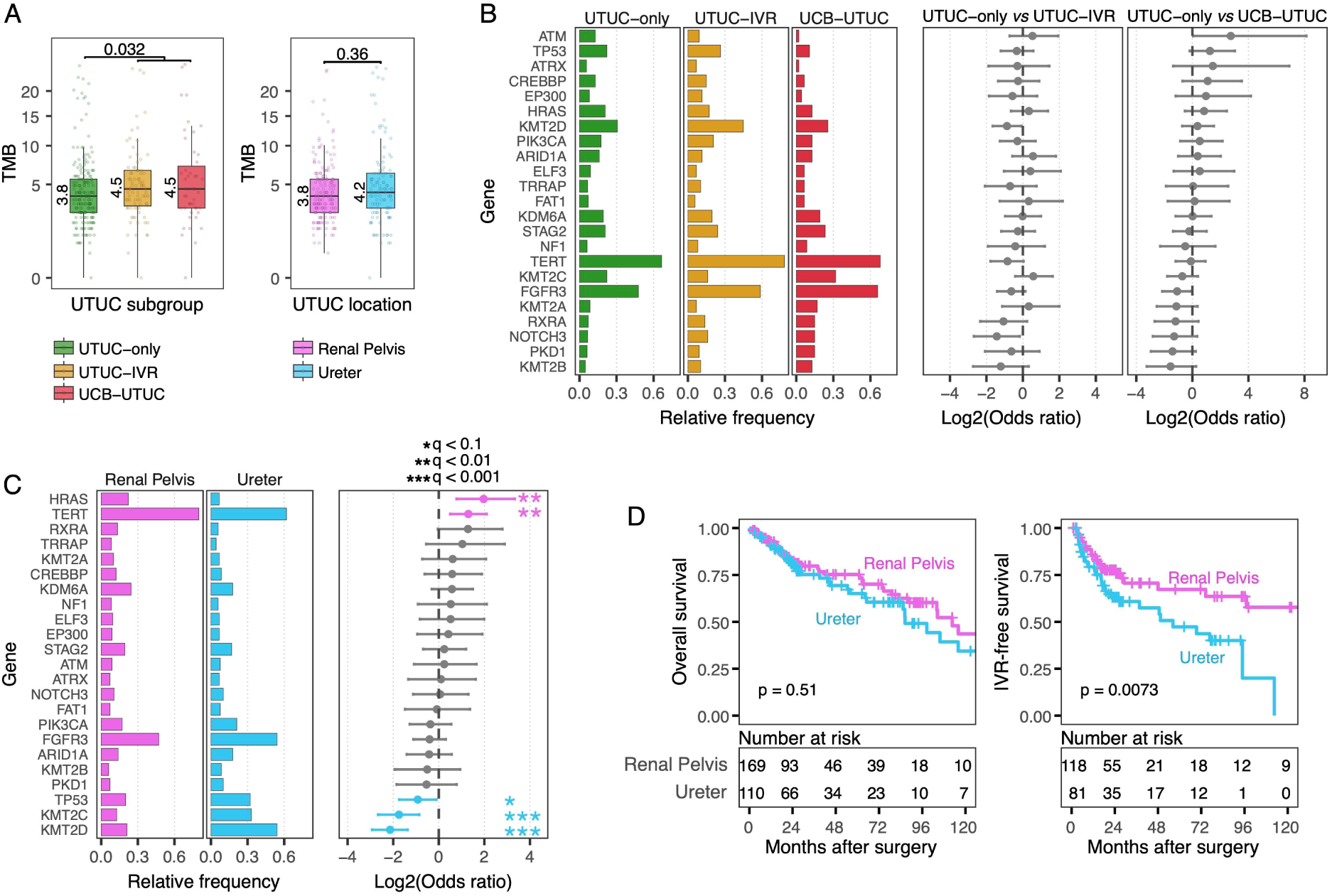
Genomic overview and differences between tumor location and between subgroups of upper urinary tract urothelial carcinoma. **A)** Tumor mutational burden (TMB) of somatic mutations, excluding hypermutated tumors, stratified by upper urinary tract urothelial carcinoma (UTUC) with a history of urothelial carcinoma of the bladder (UCB; UCB-UTUC; n=47), UTUC with intravesical recurrence (IVR; UTUC-IVR; n=83) and UTUC with neither UCB nor IVR (UTUC-only; n=206) or by UTU location: renal pelvis (n=174) and ureter (n=115). Box plots show the median, inter-quartile range (IQR: Q1–Q3) and whiskers (1.5xIQR from Q3 to the largest value within this range or 1.5xIQR from Q1 to the lowest value within this range). The two-sided Wilcoxon rank-sum test was performed for differences between UTUC-only and UCB-UTUC + UTUC-IVR, and between UTUC from the renal pelvis and ureter. Distribution of the most frequently mutated genes (≥7%) in the cohort across **B)** UTUC subgroups and **C)** UTUC location. The odds ratio between pairs of tumor groups and Benjamini-Hochberg corrected p-values (q) for two-sided Fisher’s exact test are displayed and colored accordingly. **D)** Kaplan–Meier curves for estimates of overall and IVR-free survival by UTUC location. The log-rank test was used to compare the Kaplan–Meier survival curves.

The genomic profile showed a similar distribution of mutated genes between UTUC subgroups (**Fig. 2B-C**). Similarly, time-to-event multivariate Cox regression analysis of the TMB and the most frequently mutated genes showed no correlation of genomic variables with IVR, establishing that preoperative ureteroscopy is a stronger risk factor than genomic variables (**Supplementary Fig. 4**).

Unlike UTUC subgroups, tumor location showed differential distribution of mutated genes. Somatic mutations in *TERT,* mostly in the promoter, and *HRAS* were more frequent in UTUC from the renal pelvis, while mutations in *TP53*, *KMT2C* and *KMT2D* were more frequent in UTUC from the ureter. Despite these genomic differences, overall survival was comparable between UTUC locations (**Fig. 2D**).

### Genomic subtypes in UTUC

The distribution of mutually exclusive mutated genes across all UTUC samples revealed five unsupervised hierarchical clusters (**Supplementary Fig. 5**), representing distinct genomic subtypes (GS1-5) (**Fig. 3**). GS1 (23.4%), GS3 (16.9%) and GS4 (17.1%) were enriched for *FGFR3* and *STAG2* mutations, but differed in *KDM6A*, *KMT2C,* and *KMT2D* mutations. Furthermore, GS1 was enriched for *PIK3CA* and *ELF3* mutations. GS2 tumors (16.7%) were *TP53* and *KMT2D* mutants with enrichment for wild-type *TERT* promoter mutations. GS5 (25.8%) had enrichment for *HRAS* and *KRAS* mutations.

**Figure 3.**
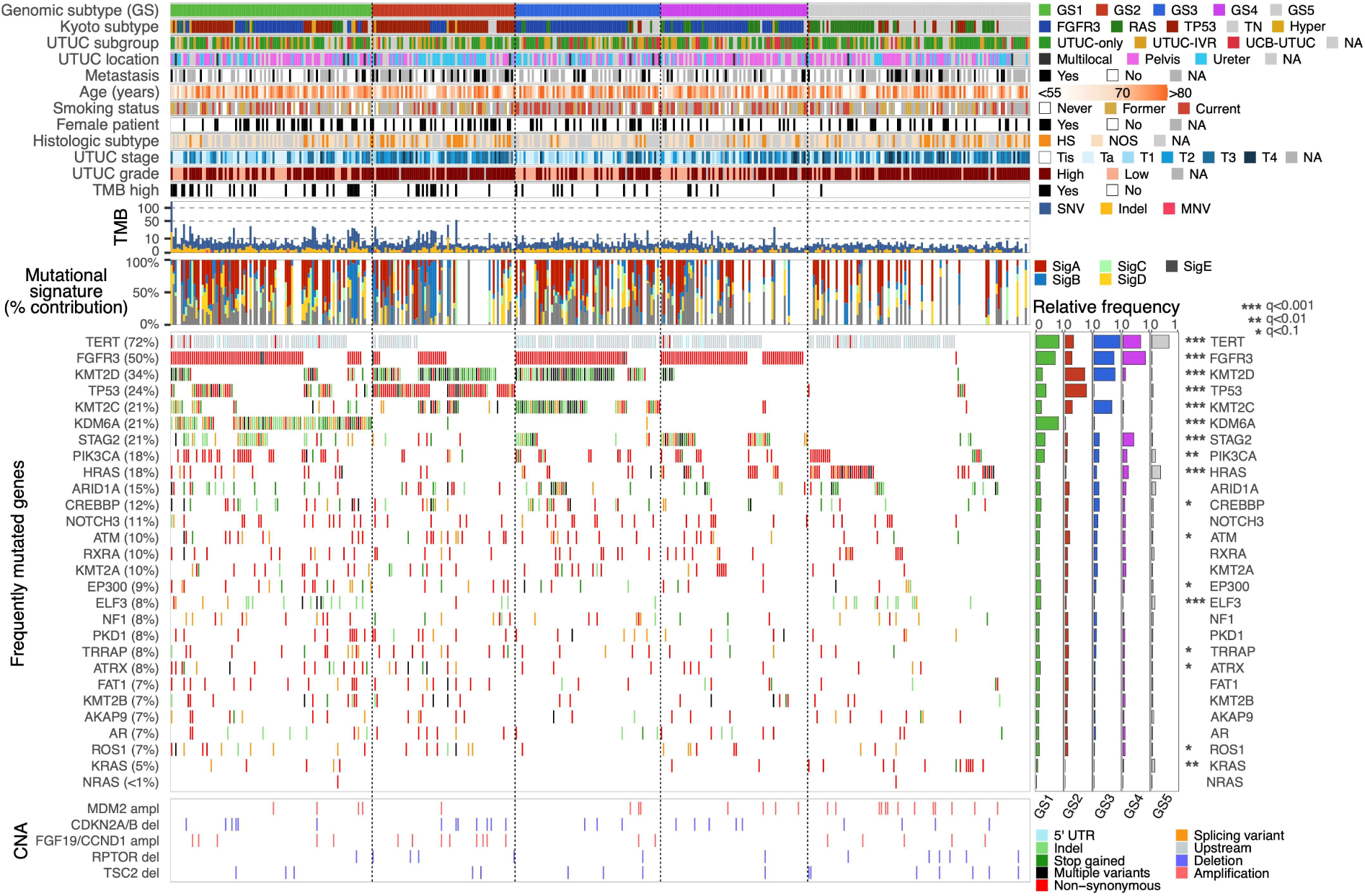
Genomic landscape of five genomic subtypes of upper urinary tract urothelial carcinoma. Tumor and patient characteristics, and genomic features of genomic subtypes of upper urinary tract urothelial carcinoma (UTUC) are displayed from top to bottom as follows: Genomic subtype of mutually exclusive mutated genes; Kyoto genomic subtype; UTUC subgroup; UTUC location; metastasis; patient age; smoking status; female patient, yes or no; histologic subtype; T stage; UTUC grade; tumor mutational burden (TMB) high tumors (>10 mutations per megabase); TMB; de novo mutational signatures (SigA-E) of tumors with ≥10 mutations; frequently mutated genes in this UTUC cohort, including *KRAS* and *NRAS,* and their frequencies across genomic subtypes applying χ^2^ test with Benjamini-Hochberg corrected p values (q); and copy number alterations (CNA) of selected genes.

We identified four *de novo* mutational signatures (**Supplementary Fig. 6, Supplementary Table 3**) related to APOBEC activity (sigA), age (sigB), aristolochic acid signature (sigC), and defective DNA mismatch repair (sigD). A fifth signature was attributed to background mutations (sigE), and all these signatures were equally distributed across genomic subtypes (**Supplementary Fig. 7A**). Further characterization of the genomic subtypes (**Supplementary Fig. 7B-K**) revealed that the distribution of UTUC subgroups, patients’ sex and age were comparable among the genomic subtypes. TMB was lowest in the GS5 subtype, while highest in the GS1 subtype (p<0.0001, Kruskal-Wallis test). UTUC from the ureter was more common in GS2 and GS3, and UTUC from the renal pelvis was enriched in GS4 (p<0.0001, χ^2^ test). Distant metastasis at follow-up and variant histology occurred more frequently in GS2 (p=0.01 and p=0.001, respectively, χ^2^ test). GS3 tumors were enriched for current smokers (p=0.003, χ^2^ test), while higher stage and high-grade tumors were frequently found in GS2 and GS5 (p<0.0001 in both, χ^2^ test).

A classifier based on a multinomial regression model was developed to assess the genomic subtypes in five external cohorts of UTUC (n=384, **Supplementary Fig. 8, Supplementary Table 4**) [10,27–30]. Despite incomplete genomic datasets for some external cohorts (e.g., lack of small insertions/deletions or unreported status of *TERT* promoter mutations), they consistently showed that GS5 had low TMB, validating our observation. We also confirmed that UTUC from the renal pelvis, high tumor stage and grade were associated with GS2. Furthermore, we confirmed that UTUC subgroups, sex and age were comparable among the genomic subtypes. In our cohort, we observed no differences in overall survival or IVR-free survival between the genomic subtypes, which was confirmed in the external cohorts. However, distant metastatic events were more common in GS2 in both our cohort and external cohorts, translating into differences in metastasis-free survival.

### Clonal evolution of intravesical recurrences

Clonality assessment of 79 matched UTUC-IVR cases found 92% (73/79) of IVR to be clonally related to UTUC, which was accompanied by an increment in the TMB (**Fig. 4A-B**; p=0.027, two-sided Wilcoxon signed-rank test). In more than half of the ‘non-clonal’ cases (4/6), clonality assessment was limited due to a low number (≤2) or lack of mutations in the matched UTUC-IVR (**Supplementary Table 5**), suggesting that the proportion of clonality might have been higher than what we report here. In 70% (55/79) of paired UTUC-IVR cases, mutations in either the *TERT* promoter, *FGFR3* or *HRAS* were identified (**Fig. 4C**).

**Figure 4.**
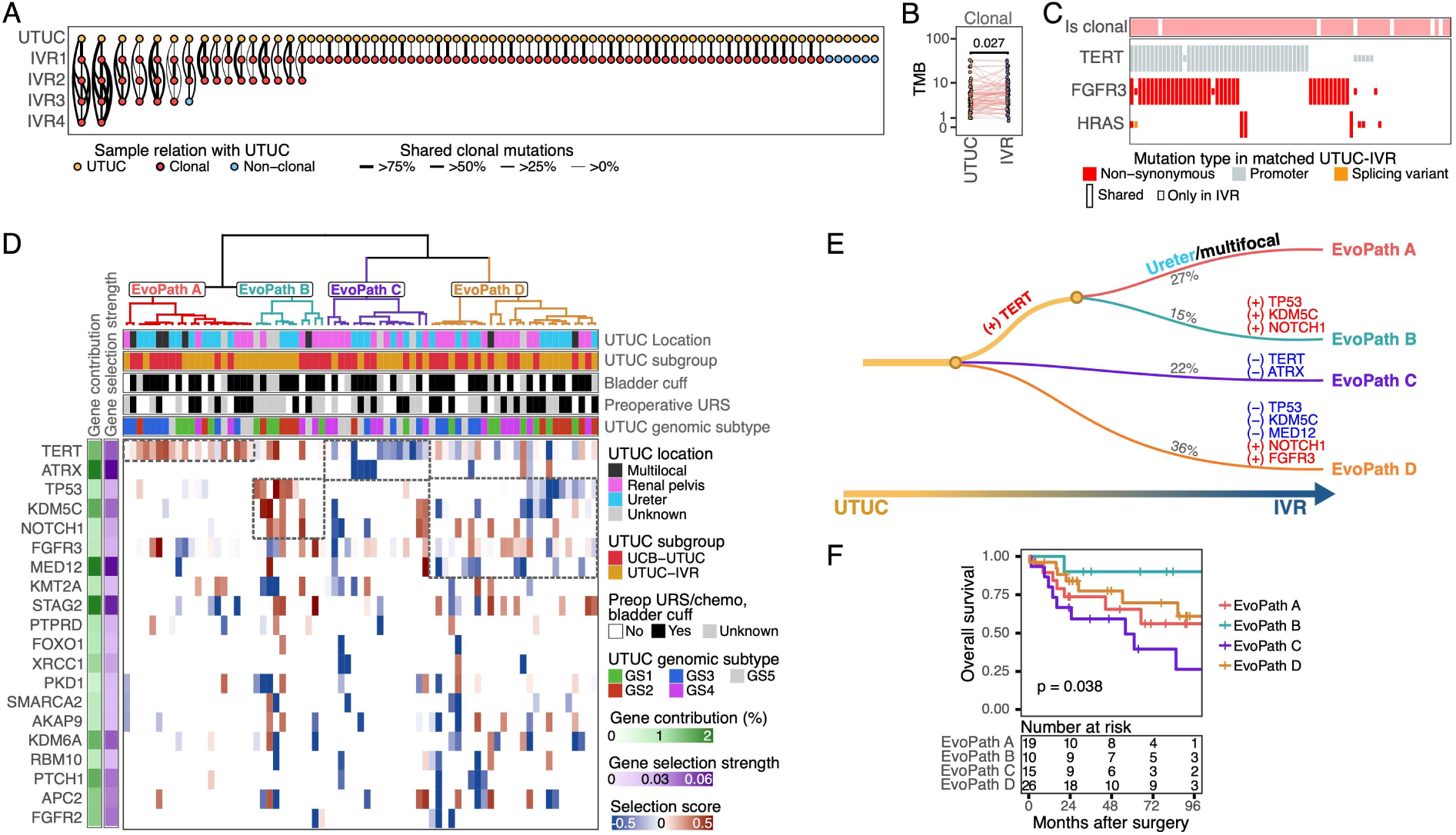
The clonal evolution of upper urinary tract urothelial carcinoma and intravesical recurrences. **A)** Evolutionary trees of all upper urinary tract urothelial carcinomas (UTUC) and their corresponding intravesical recurrences (IVR) per patient. Clonal relations between UTUC and IVRs (IVR1-4) are represented by connecting lines. **B)** Comparison of the tumor mutational burden (TMB) between UTUC and IVR for clonally related cases using the average TMB for multiple IVRs (two-sided Wilcoxon signed-rank test). **C)** Hotspot mutations in the *TERT* promoter, *FGFR3* and *HRAS* in UTUC and IVR. **D)** Hierarchical clustering of the selection score in genes mutated in at least two patients. Only genes with the top 20 strongest selection scores and with the highest contribution to variability across cases are displayed. **E)** Graphical representation of the four clusters from (D) into evolutionary paths (EvoPath A-D) of IVR. **F)** Kaplan-Meier survival curves of EvoPaths. P-value was estimated with the log-rank test.

All clonally related UTUC-IVR (n=73) were further analyzed to define the evolution of UTUC towards IVR. Hierarchical clustering of the selection scores of mutated genes revealed four evolutionary paths (EvoPaths A-D; **Fig. 4D-E**), which were not influenced by a history of UCB or treatment such as bladder cuff excision or preoperative ureteroscopy. The genomic subtypes were differentially distributed across the evolutionary paths (p=0.055, χ^2^ test), with GS2 underrepresented in EvoPath A and C. EvoPath A (27%), driven by positive selection of *TERT* promoter mutations, was favored by multilocal and ureteral tumors (p=0.042, one-sided Fisher’s exact test). In addition to *TERT,* EvoPath B (15%) was also driven by positive selection of mutant *TP53*, *KMD5C,* and *NOTCH1*. EvoPath C (22%) was characterized by negative selection of *TERT* and *ATRX* mutant genes, and these patients had the poorest overall survival compared to other EvoPaths (**Fig. 4F**). Tumors in EvoPath D (36%) followed a more complex path involving positive and negative selection of several genes, including *TP53, KMD5C*, *NOTCH1, MED12 and FGFR3* mutations.

## Discussion

UTUC is a relatively rare form of UC with a high risk of recurrence in the bladder following surgery, for which the molecular characteristics are not well understood. In this study, we aimed to characterize the genomic landscape of 414 UTUC patients treated with radical surgery and identify molecular markers of IVR. In-depth analysis of 79 patients with paired UTUC-IVR further refined our results and delineated the clonal evolution of IVR.

Our analysis failed to validate previous studies that have linked mutations in *FGFR3*, *KDM6A,* and *CCND1* with increased risk of IVR [19]. In our study, a diagnostic ureteroscopy prior to RNU was a very strong risk factor for IVR, which is in line with multiple other studies [31,32]. This suggests that alternatives for the diagnosis of UTUC, such as liquid biopsies and percutaneous needle-core biopsies [32–35], need further investigation to significantly reduce the risk of IVR after UTUC.

Further analysis of the molecular data identified five genomic subtypes of UTUC, but despite their unique clinical characteristics, no differences were observed in the risk of IVR. These subtypes only partially overlapped with the previously defined genomic subtypes of UTUC [30], offering more granularity for *FGFR3*-mutated tumors, which affected 50% of UTUC patients. Although the clinical benefit of these genomic subtypes is not yet clear, their distinct biology may translate into differential responses to therapeutic agents, especially for the three genomic subtypes carrying *FGFR3* mutations. For instance, GS3 carries *KMT2C/D* mutations, which have been shown to upregulate epidermal growth factor receptor (EGFR) signaling and confer vulnerability to EGFR inhibitors [36]. However, since GS3 harbors *FGFR3* mutations as well, combination therapy with an anti-FGFR may potentiate therapeutic response in these patients [37].

A clonal relationship between UTUC and paired IVR was observed in 92% of patients, which is in line with other studies reporting 73-100% [18,19]. This result, together with *TERT* promoter, *FGFR3* and *HRAS* mutations in over two-thirds of cases, underscores the potential of surveillance by molecular testing. Urine assays that include these three genes, among other markers, have shown important diagnostic accuracy [38,39], which could accelerate their clinical implementation for non-invasive surveillance of patients after surgery for UTUC.

UTUC develops differently across locations, showing different sets of driver genes [8,30]. These differences are also reflected in the distinct evolutionary paths of UTUC towards the bladder, which were not linked to previous clinical interventions or history of UCB. We speculate that positive selection of *FGFR3* mutations during the evolution of IVR suggests potential benefit of an anti-FGFR to reduce the risk of IVR; however, negative selection of *FGFR3* mutations in some cases anticipates a more complex scenario that may be tackled with combination therapy.

Limitations of our study are the use of targeted DNA-sequencing, restricting our analysis to 571 genes and heterogeneity between the included cohorts. The prospective cohort had a shorter follow-up and excluded patients with a history of UCB, which may have impacted data interpretation.

## Conclusion

This study advances the molecular characterization of UTUC and reinforces the central role that FGFR-directed therapies may play in shaping future treatments, potentially in combination with other therapeutic agents. Our study highlights the need for new (less-)non-invasive approaches for the diagnosis of UTUC to reduce the risk of IVR. Similarly, the high clonality rate of IVR with UTUC encourages further investigation into the potential of urine-based molecular assays for surveillance of patients undergoing radical surgery for UTUC.

## Data Availability

Raw sequencing data have been deposited in EGA (EGAS50000001805) and a summary of the clinical data and processed genomic data that support the findings of this study are available in the Supplementary Tables.

## Acknowledgements

We thank all local principal investigators and the nurses of all contributing centers for their help with patient recruitment. We are particularly grateful to all participating patients and their families.

## Author Contributions

Conceptualization: AH, BH-I, YA, JLB;

Methodology: JAN-G, VB, TvD, JH, AH, BH-I, YA, JLB;

Data analysis: JAN-G, JP, JMP, IB;

Data interpretation: JAN-G, HJGvdW, JH, MFL, YA, JLB;

Clinical data collection and interpretation: VB, TvD, JH, TW, GJLHvL, MFL, FP-P, EG-G, RML, LG-M, ME, RS, DS, IGM, JDPC, MJL, MA, EM-R, AH, BH-I, YA, JLB.

Writing – Original Draft: JAN-G, JLB;

Writing – Review & Editing: All authors.

All authors read and approved the final manuscript.

## Data availability

Raw sequencing data have been deposited in EGA (EGAS50000001805) and will be released when the paper is published. Data from external cohorts are freely accessible at https://www.cbioportal.org/. Pre-processed data from the Tokyo cohort were kindly provided by Seishi Ogawa and Yoichi Fujii [30].

## Code availability

The genomic subtype classifier is available as an R package at https://github.com/erasmus-ur/UTUCclassifyGS

