## Supplementary Fig. for "Genomic characterization of upper urinary tract urothelial carcinoma and clonal evolution of intravesical recurrences"

#### Supplementary Methods

##### Patient cohort and study design

Patients diagnosed with upper tract urothelial carcinoma (UTUC) between 2002 and 2020 were included in an international multi-institutional cohort. Patient samples in the context of the REBACARE trial were collected prospectively in 18 Dutch hospitals from 2017-2020 [1], while the others were retrospectively collected in Germany and Spain. The study protocol was approved by the medical ethics review board of the Erasmus University Medical Center, the Netherlands (METC 2017-227, NL60919.078.17), and obtained enforceability permission for all other sites. Written informed consent was obtained from all participants prior to inclusion in the study. The study population consisted of 448 adult patients with UTUC without distant metastatic disease whose clinical data were collected using a standardized electronic data capture tool. DNA from tumor and normal-adjacent urothelium or renal tissue was extracted from archived Formalin-Fixed Paraffin-Embedded (FFPE) blocks. In patients from the prospective cohort, germline DNA was isolated from buccal swabs. DNA extraction was performed following the same protocol and kits in all the institutes. All cases from the retrospective cohorts were centrally reviewed by two uropathologists (AH, VB) according to the latest Tumor Nodal Metastasis (TNM) staging manual of the UICC 8th edition, 2017, and the World Health Organization (WHO) 2022 classification for tumors of the genitourinary tract. For the prospective cohort, pathology reports from the local medical centers where patients were surgically treated were used. IVR was defined as a histologically proven urothelial carcinoma of the bladder.

**DNA-sequencing and analysis**

Tumor DNA, together with matched normal samples, was centrally collected (Institut Curie, France) for targeted next-generation DNA sequencing using a custom panel called DRAGON (Detection of Relevant Alterations in Genes involved in Oncogenetics by NGS), commercially available as SureSelect CD Curie CGP by Agilent [2]. This panel encompasses 571 oncology-related genes spanning 2.87 megabases, serving diagnostic, prognostic, therapeutic, and predisposition purposes. The DRAGON pipeline has been successfully applied and extensively described in other studies to assess somatic mutations and copy number alterations [2, 3]. In short, next-generation sequencing libraries were prepared using an Agilent SureSelect XT-HS kit, and sequencing was performed in NovaSeq (Illumina). Reads were mapped using *BWA mem* (v0.7.15) on the Human reference genome (hg19 assembly) using default parameters. Copy number alterations were estimated with a modified version of the facets R package (v0.6.0) and variant calling of single-nucleotide variants (SNVs) and small insertion/deletions (indels) was performed using Varscan2 (v2.4.3). Annotations were provided by ANNOVAR (04/16/2018 version) and supplemented with the hgvs R package (v1.2.5). Of the initial 1099 tumor and normal DNA-sequenced samples, 1005 with sufficient coverage were kept for processing. Somatic mutations were retained if the allele frequency in the tumor was >0.05 with a minimum read depth >30x, and allele frequency in normal was <0.05 with a minimum depth of 20x. In the case of known somatic hotspot mutations (in *TERT*, *FGFR3*, *HRAS*, *PI3KCA*, *TP53*), the allele frequency in normal samples was not considered. Due to variability in sample quality because of FFPE preservation, some samples presented an extremely large number of mutations. These false positive somatic mutations followed an exponential distribution that we used to set a higher allele frequency cut-off for tumors with long exponential tails. Using the distribution of allele frequencies in the range [0.05, 0.20] with intervals of 0.005, the decay towards $y_f$ was estimated and used as key parameter to define the allele frequency cut-offs at 0.05, 0.10, 0.15, 0.20, 0.25, and 0.30 when $y_f$ was <-1, <0.22, <1.2, <4.5, <7 or <39, respectively. In case of $y_f$>39, the sample was considered of very bad quality and excluded from further analysis. For good-quality samples but insufficient mutations, fitting the exponential function to disperse data points was unsuccessful and the cut-offs for these samples were kept at 0.05. Manual inspection for quality check was used as a final step to either include or exclude samples. In case of multiple samples from the same patient, mutations with allele frequency > 0.01 were rescued if mutations passed all the filters in at least one of the other samples. This strategy ensures that clonal mutations with low frequency are considered for paired UTUC-intravesical recurrence (IVR) analysis. Thus, a total of 518 (414 UTUC + 104 IVR) tumor samples with their corresponding 414 paired normal samples were retained for downstream analysis.

Tumor mutational burden (TMB) was calculated as the number of all mutations divided by 2.87, which is the coverage of the DRAGON panel. *De novo* mutational signature calling was performed on UTUC samples with ≥10 SNVs. Samples with few mutations may not give reliable results and were excluded. The NMF v0.27 R package was used to define the number of *de novo* signatures and MutationalPatterns v3.14 was applied to extract such signatures.

Following Fujii *et al*.[4], we defined the Kyoto molecular subtypes of UTUC. In this cohort, we considered a sample hyper-mutated if TMB>28.
For the novo genomic subtypes, the ten most significant mutually exclusive genes (*TERT*, *FGFR3*, *KMT2D*, *KMT2C*, *TP53*, *KDM6A*, *HRAS*, *KRAS*, *STAG2* and *ELF3*) were selected for hierarchical clustering. A solution of five clusters was used to represent genomic subtypes, and a multinomial regression model, using nnet v7.3-20, was developed for a single-sample classifier based on the absence or presence of mutations.

**Clonality assessment and evolution in UTUC-IVR paired samples**

The clonal relationship between UTUC and IVR from the same patient was evaluated with the clonal relatedness test based on Neyman-Pearson Lemma test and the conditional maximum likelihood estimate developed by Ostrovnaya *et al*. [5, 6]. To apply this test, a mutation reference data set was constructed by estimating the relative frequencies of SNVs and indels present in the samples of all UTUC patients. For patients with multiple samples, only one mutation per patient was allowed. Note that hotspot mutations would have high frequencies and rare mutations would have the lowest frequencies (1/414=0.0024), which the clonal relatedness test uses as probabilities of finding a given mutation in a sample. The test applies a pairwise comparison of UTUC-IVR pairs with 100,000 simulations and p-values are corrected for multiple testing with the Benjamini-Hochberg method.

Clonal evolution was interrogated in samples that were clonally related (n = 73). In case of multiple IVRs, the first IVR (IVR1) was used. Synonymous and non-coding mutations, except promoter mutations (downstream, upstream or UTR), were excluded as the impact of these mutations in genes is not well documented. The change in allele frequency (AF) of specific mutations between IVR and UTUC was calculated as $1 - e^{AF\_IVR}/e^{AF\_UTUC}$. Positive values represent an increase in the AF of mutations, while negative values represent a decrease. We considered these values as a proxy for selection because these mutations can be beneficial or detrimental to subclonal populations when colonizing new niches (the bladder), which is reflected in an increase or decrease in the AF, respectively. Alternatively, copy number changes may also alter the AF of these mutations, indicating their benefit or not in the new environment. Some genes may have more than one mutation, and in this case, a mean value was calculated that we called the gene selection score. The selection scores of genes mutated in at least two patients were used for hierarchical clustering with ConsensusClusterPlus v1.68.0.

### Supplementary Figures

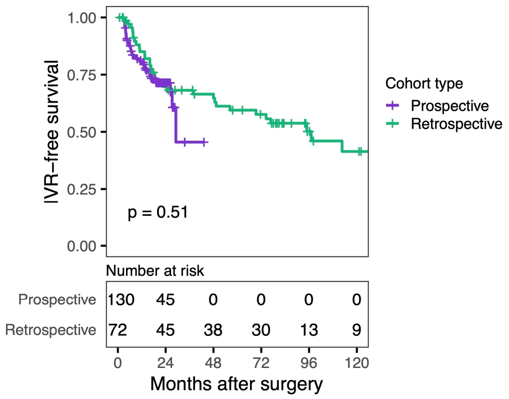


**Supplementary Figure 1. Intravesical recurrence free survival comparison between the prospective and retrospective cohorts.** Kaplan–Meier curves for estimates of intravesical recurrence (IVR) among cohorts were compared using the log-rank test.

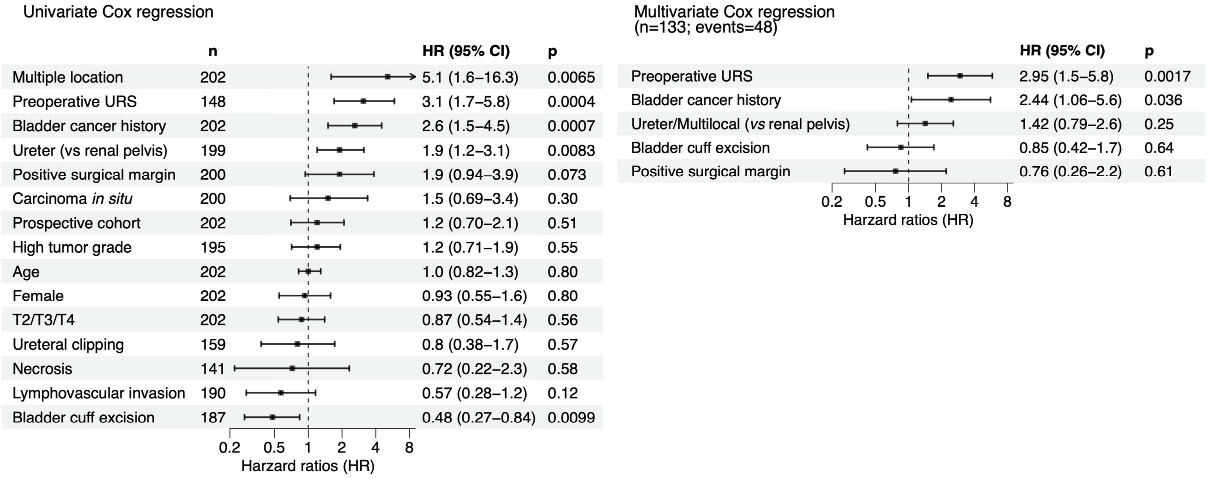


**Supplementary Figure 2. Univariate and multivariate Cox regression analysis for clinicopathological characteristics corresponding to the risk of intravesical recurrence.** The Cox proportional hazards regression analysis was applied to time to intravesical recurrences (IVR) after surgery. The prospective cohort corresponds to patients from one institution (Erasmus MC, the Netherlands). Multiple location tumors occurring in both the renal pelvis and ureter were compared against those occurring in only one location. All variables were categorized except for age, for which continuous values were used. Arrows indicate when values extend beyond the displayed scale. For multivariate analysis (n=133 with complete data), only variables with p<0.1 in the univariate analysis were used and due to the low number of cases, multiple locations and ureter were combined. Hazard ratios (HR) and 95% confidence intervals (CI) are displayed.

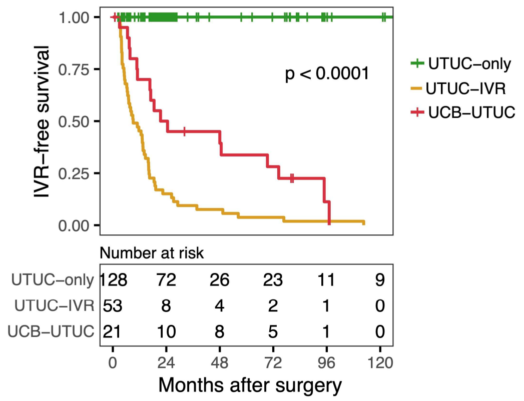


**Supplementary Figure 3. Intravesical recurrence-free survival comparison between UTUC subgroups.** Kaplan–Meier curves for estimates of intravesical recurrence (IVR) among UTUC subgroups were compared using the log-rank test.

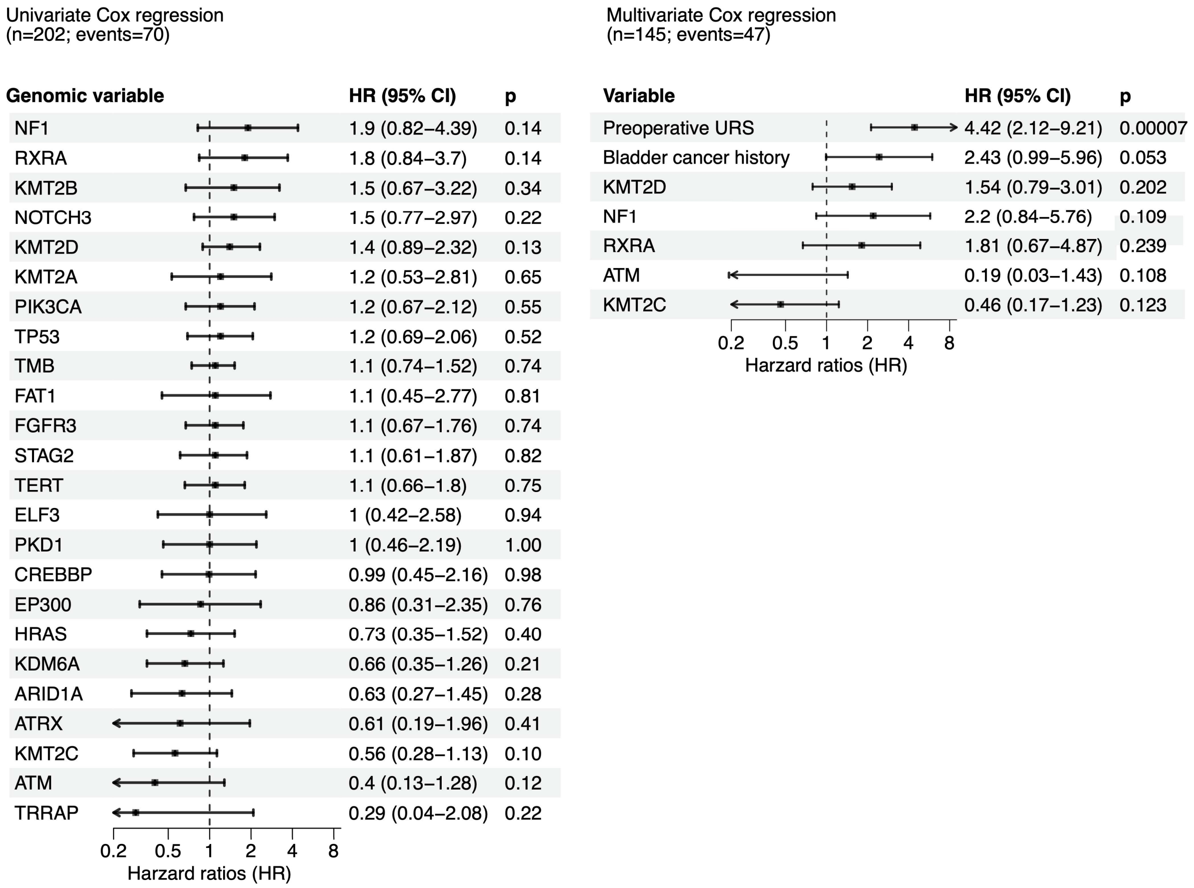


**Supplementary Figure 4. Multivariate Cox regression analysis for genomic features corresponding to the risk of intravesical recurrences.** The Cox proportional hazards regression analysis was applied to time to intravesical recurrences (IVR) after surgery. The most frequently mutated genes and tumor mutational burden (TMB) were assessed. All variables were categorized (mutant or wildtype) except for TMB, for which continuous values were used. Arrows indicate when values extend beyond the displayed scale. For multivariate analysis, only variables with p<0.2 in the univariate analysis were used, together with significantly associated clinicopathological variables with IVR (preoperative ureteroscopy and bladder cancer history). Hazard ratios (HR) and 95% confidence intervals (CI) are displayed.

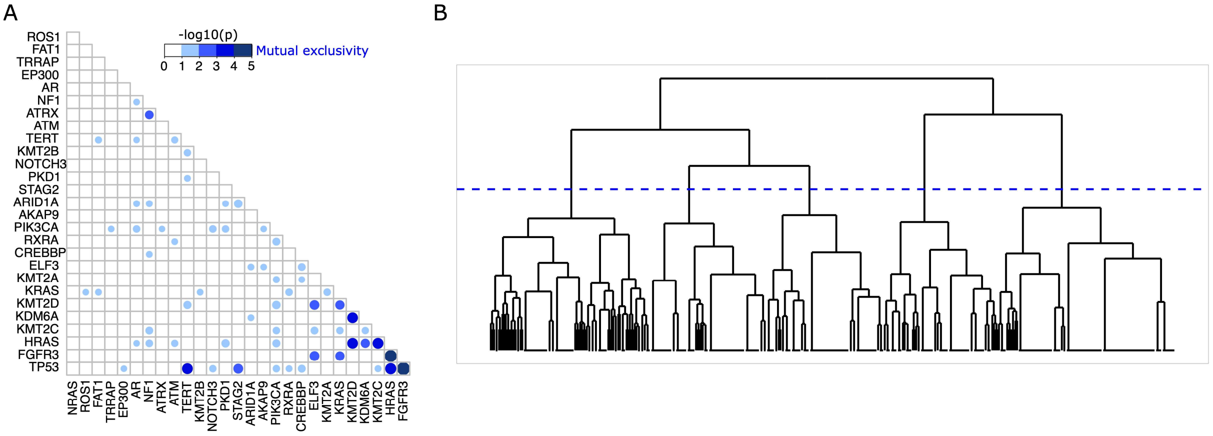


**Supplementary Figure 5. Mutually exclusive genes and genomic clusters. A)** Mutual exclusivity test for highly mutated genes. **B)** Hierarchical clustering using Euclidean distance and ward.D2 method for genes with the 10 most significant p-values. The horizontal blue dashed line indicates the cut-off for selecting five clusters.

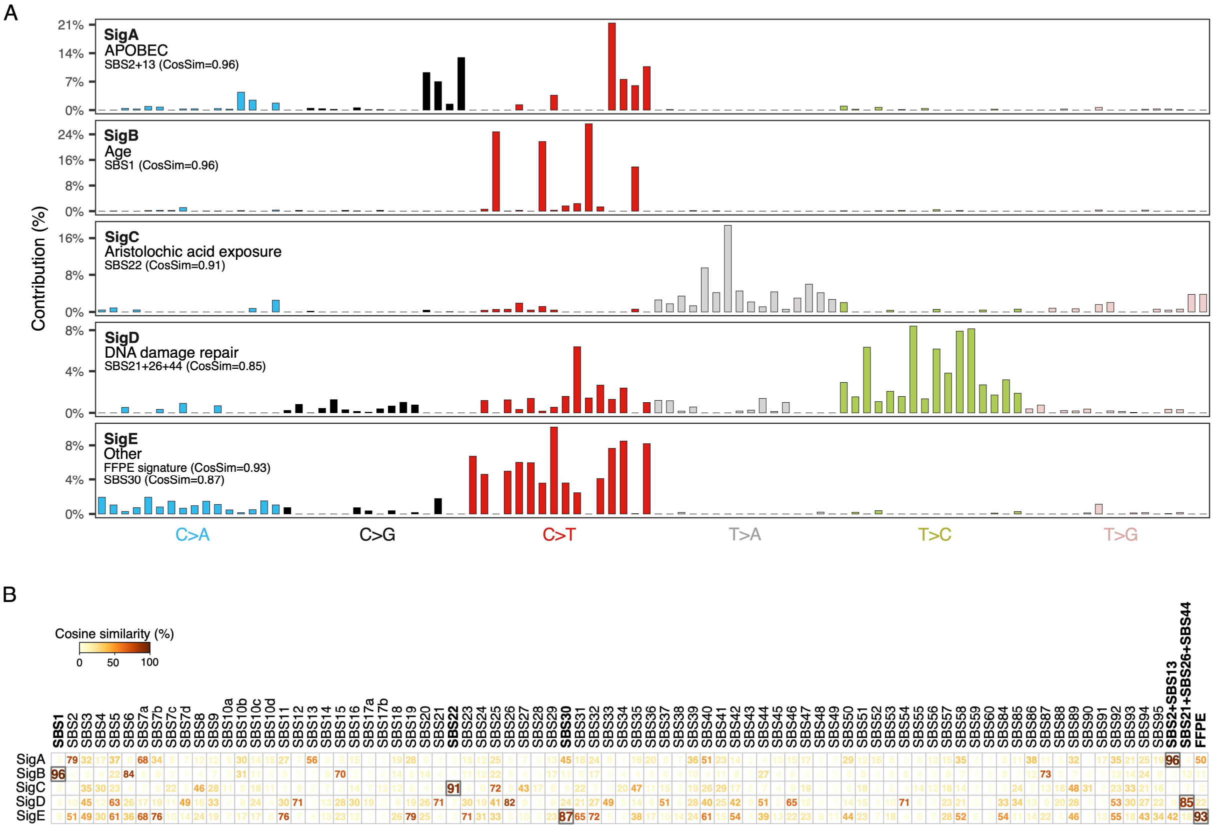


**Supplementary Figure 6. *De novo* mutational signatures in upper urinary tract urothelial carcinoma. A)** *De novo* mutational signatures performed in upper urinary tract urothelial carcinoma (UTUC) samples with ≥10 single-nucleotide variants (n=228), and their highest cosine similarity (CosSim) with COSMIC signatures v3.3. **B)** Cosine similarity of the five de novo mutational signatures with established COSMIC signatures v3.3 and FFPE damage signature.

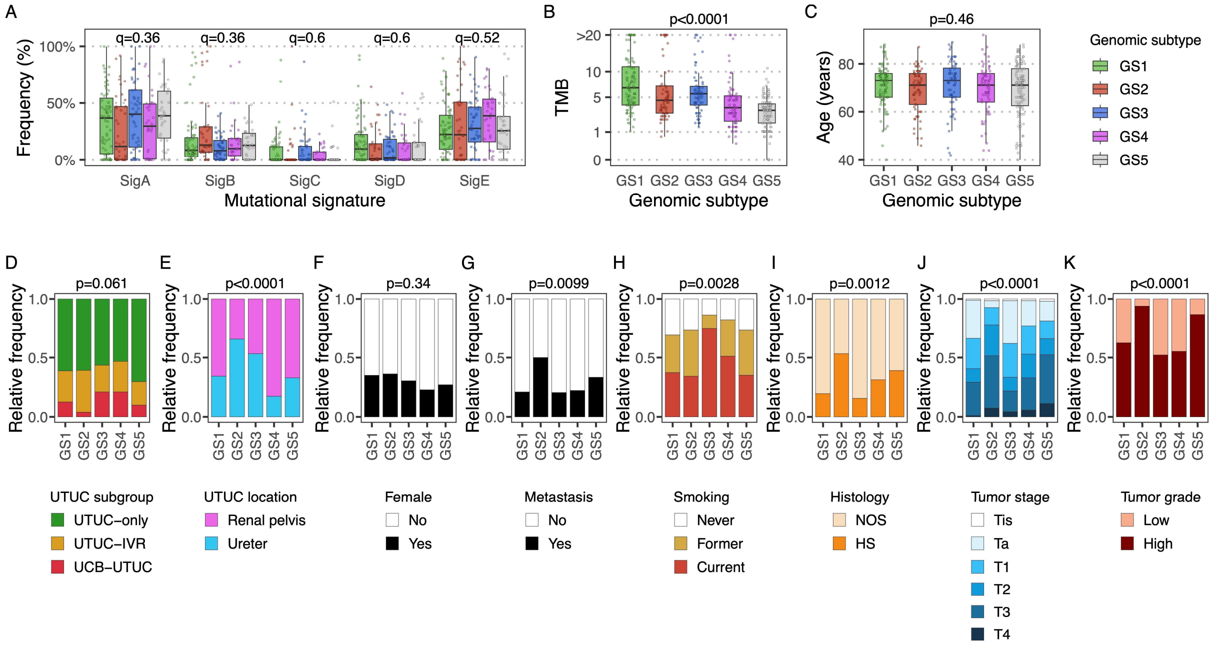


**Supplementary Figure 7. Molecular and clinicopathological characteristics of the Genomic Subtypes (GS). A)** *de Novo* Mutational signatures; **B)** tumor mutational burden (TMB); **C)** patient age; **D)** upper urinary tract urothelial carcinoma (UTUC) subgroup; **E)** UTUC location; **F)** female patients; **G)** metastatic event; **H)** smoking history; **I)** histology; **J)** tumor stage; **K)** tumor grade. The Kruskal-Wallis test was applied for continuous values and the $\chi^2$ test for categorical variables. Benjamini-Hochberg correction for multiple testing was applied in A).

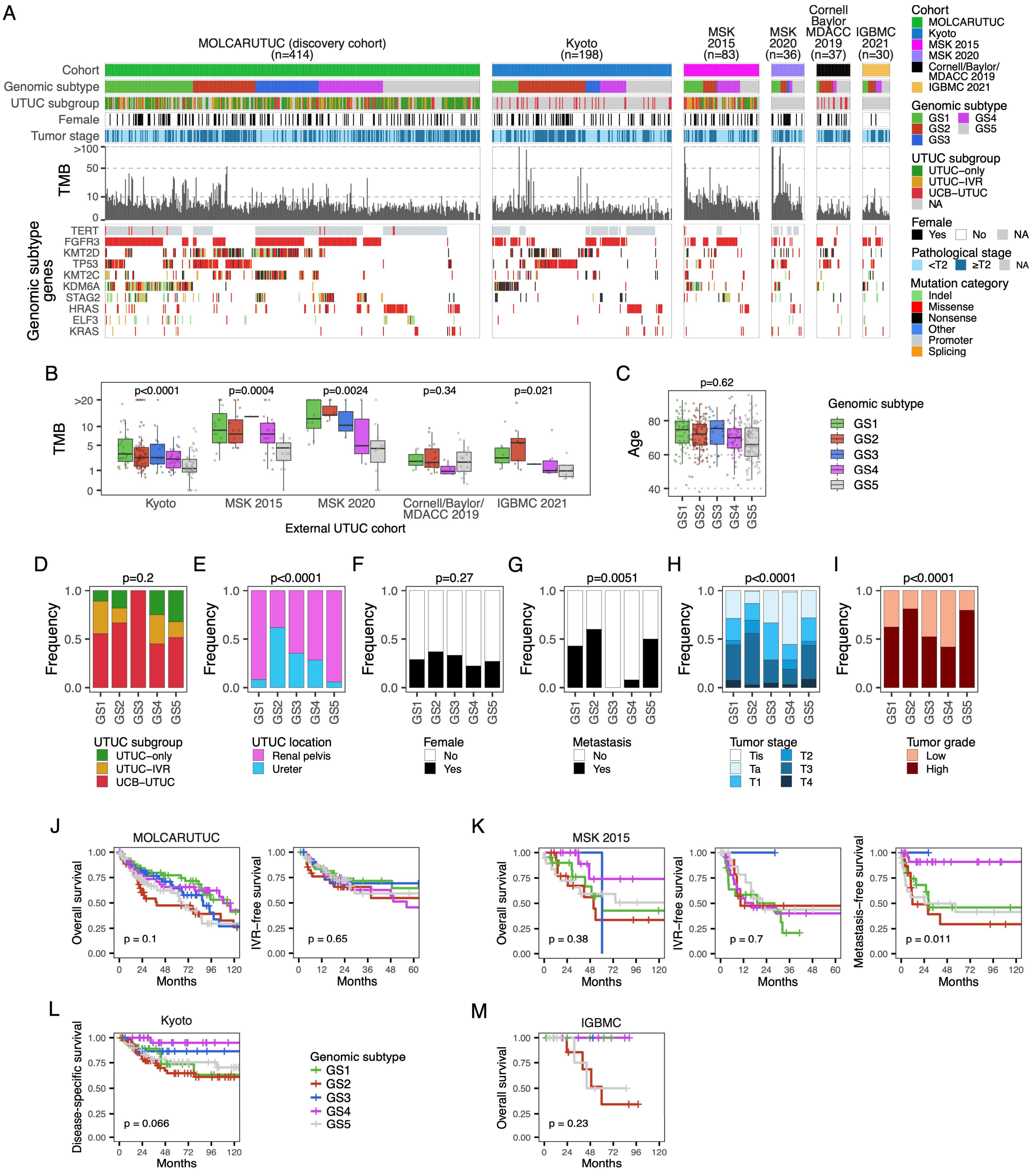


**Supplementary Figure 8. Genomic subtypes of upper urinary tract urothelial carcinoma across external cohorts. A)** Clinical and molecular characteristics of external cohorts stratified by genomic subtypes. The discovery cohort (MOLCARUTUC) is included for reference. From top to bottom: cohort; genomic subtype of upper urinary tract urothelial carcinoma (UTUC); UTUC subgroup; female patients; tumor stage; tumor mutational burden (TMB); somatic mutations of genes that define the genomic subtypes. **B)** TMB of somatic mutations across external cohorts and stratified by genomic subtype**.** The Kruskal-Wallis test was applied. **C)** Age of patient in external cohorts combined. The Kruskal-Wallis test was applied. $\chi^2$ test was applied for **D)** UTUC subgroup, **E)** UTUC location, **F)** female patients, **G)** metastatic event, **H)** tumor stage and **I)** tumor grade. Kaplan-Meier survival curves of genomic subtypes in the **J)** discovery and **K-M)** external cohorts. P values were estimated with the log-rank test. Due to insufficient data availability for external cohorts, variables in C)-I) were combined from all external cohorts.
